# Leveraging the genetics of human face shape boosts the discovery of orofacial cleft risk loci

**DOI:** 10.64898/2026.01.30.26345139

**Authors:** Noah Herrick, Seppe Goovaerts, Alexandra Manchel, Myoung Keun Lee, Xinyi Zhang, Amy Davies, Jenna C Carlson, Elizabeth J Leslie-Clarkson, Sarah J Lewis, Mary L Marazita, Justin Cotney, Peter Claes, John R Shaffer, Seth M Weinberg

## Abstract

Several lines of evidence suggest that normal-range facial features and nonsyndromic orofacial clefts (OFCs) exhibit a shared genetic basis. Approaches designed to leverage this relationship hold the possibility of revealing new OFC risk loci by boosting discovery power. To test this idea, we applied a pleiotropy-informed GWAS method (cFDR-GWAS) with summary statistics from large, independent European GWASs of normal facial shape (n=4,680; n=3,566) and nonsyndromic cleft lip with or without cleft palate (nsCL/P, n=3,969). The cFDR approach identified 21 independent genomic loci significantly associated with nsCL/P, providing further evidence of the interconnected genetic architecture between these traits. The five original nsCL/P GWAS signals were detected and joined by nine additional loci previously implicated in other OFC association studies. The remaining seven loci represent new nsCL/P genomic regions, and three of these replicated (P < 0.05) in an independent nsCL/P cohort: *ASPSCR1, MSX2*, and *RALYL*. A relaxed 10% cFDR-GWAS threshold identified 15 more independent loci with comparable effect sizes to those detected at the strict 5% threshold, two of which replicated: *FHOD3* and *SMARCA2*. Gene expression patterns in major cell types and spatial transcriptomics data highlighted our gene candidates’ roles in craniofacial development. In conclusion, the application of an empirical Bayesian strategy to draw on association signals from genetically related traits can boost the power to identify and prioritize OFC risk loci missed by agnostic gene mapping approaches. These results hold promise that the cFDR-GWAS approach may be able to enhance our understanding of the genetic architecture of other structural birth defects.

## INTRODUCTION

Orofacial clefts (OFCs) are among the most common and recognizable congenital malformations affecting humans, with an average global incidence estimated at 1/700 live births^1^. OFCs can take several distinct forms with the large majority of cases presenting as isolated malformations and the remainder as non-isolated/syndromic. Affected individuals typically require multiple surgeries and ongoing care from a range of specialists. Moreover, OFCs are associated with increased morbidity across the lifespan, and there is some evidence for an elevated risk for certain cancers^2-5^. Despite the relatively high incidence and the individual and public health burdens, the underlying causes behind OFCs remain poorly understood. This is due in part to the trait’s complex and multifactorial etiology^6^ involving a heterogeneous array of genetic and environmental risk factors and their interactions. While genome scans have identified over 50 loci associated with different forms of nonsyndromic OFCs^7-18^, these associations explain only a fraction of the heritability. It is therefore likely that many genetic risk variants have yet to be detected, particularly if their phenotypic effects are small, they act as modifiers, or their effects are influenced by environmental factors. Very large sample sizes are often needed to detect such effects, but unlike ubiquitous traits such as BMI or blood pressure, this is not possible for genetic studies of congenital malformations such as OFCs. Therefore, analytical approaches are needed that can increase our chances of finding such difficult to detect risk variants and thus help explain the heritability of these common congenital anomalies.

One method for boosting the discovery of common variants for complex traits is to leverage their shared genetic architecture with other closely related traits. First proposed to investigate psychiatric disorders^19^ and reintroduced in a recent review^20^, the pleiotropy-informed conditional false discovery rate (cFDR) approach works by testing the association of genetic variants with a target trait while controlling for their association with a second related trait. In other words, the method generalizes empirical Bayes^21^ to assign conditional FDR values to genetic variants that show prior association with a closely related trait. This increases power to detect true non-null effects present at some level in both traits. Additionally, this effectively reduces the FDR for the target trait, potentially uncovering novel loci that might remain undetected in a traditional GWAS approach for a given sample size.

In this study, we apply the conditional FDR approach to re-examine the genetic basis of nonsyndromic cleft lip with or without cleft palate (nsCL/P), the most common subtype of OFC. For the conditioning trait we focus on human face shape. Since 2012, multiple GWAS in different populations have identified over 300 loci impacting different human facial features (reviewed by Naqvi et al., 2022)^22^. There is now ample evidence that many of the genes involved in human facial shape have also been implicated in nonsyndromic OFCs. For example, our team found that genes nominated at GWAS signals for face shape showed significant enrichment for biological processes and phenotypes linked to clefting^23,24.^ We also directly tested lead SNPs from prior nsCL/P GWASs and showed that they have a significant effect on human facial shape^25^. Moreover, we quantified a risk endophenotype from three-dimensional photos of unaffected parents of children with nsCL/P, projected that endophenotype onto a cohort of normal-range faces for GWAS, and identified 29 SNPs significantly associated with aspects of the endophenotype in the philtrum, nose, mandible, and upper face^26^. The link between face shape and clefting predates the era of modern gene mapping and has been actively investigated since the 1960s (reviewed by Weinberg, 2022)^27^. This link is premised on the idea that embryonic facial shape is itself a risk factor for clefting because altering the position and morphology of the rapidly growing facial prominences could impede the union of adjacent tissues in the developing face. Thus, conditioning on face shape may reveal the genes and pathways involved in conferring that risk and could offer insights into an important potential mechanism for OFCs.

## RESULTS

### OFC loci are enriched for facial genetic associations

To highlight the enrichment of nsCL/P-associated SNPs in facial morphology GWAS results compared to other non-craniofacial complex trait GWAS results, we calculated the ratio of SNPs nominally associated with nsCL/P (P < 0.05) within the subset of SNPs also showing nominal association with a second trait, compared to all SNPs genome-wide. An enrichment of nsCL/P-associated SNPs was evident among those associated with facial shape traits but not to other complex traits (Figure 1a), further supporting the overlapping genetic architecture described in previous work on this intersection of OFCs and face shape. This type of enrichment aligns with the Bayesian logic underlying the cFDR framework used in subsequent analyses. Thus, we also compared these patterns to genome-wide Spearman correlations—a complementary metric of genetic correlation suitable for multivariate GWAS data. This comparison indicated that as enrichments increased between traits so did genomic correlations, starting with low enrichments and low correlations between nsCL/P and several non-craniofacial traits^28-34^ and trending toward high enrichments and high correlations between nsCL/P and facial shape. These findings support the idea that when two traits are genetically correlated, such as nsCL/P and facial shape, the presence of association with one trait increases the likelihood that a given SNP is also involved in the other.

**Figure 1.**
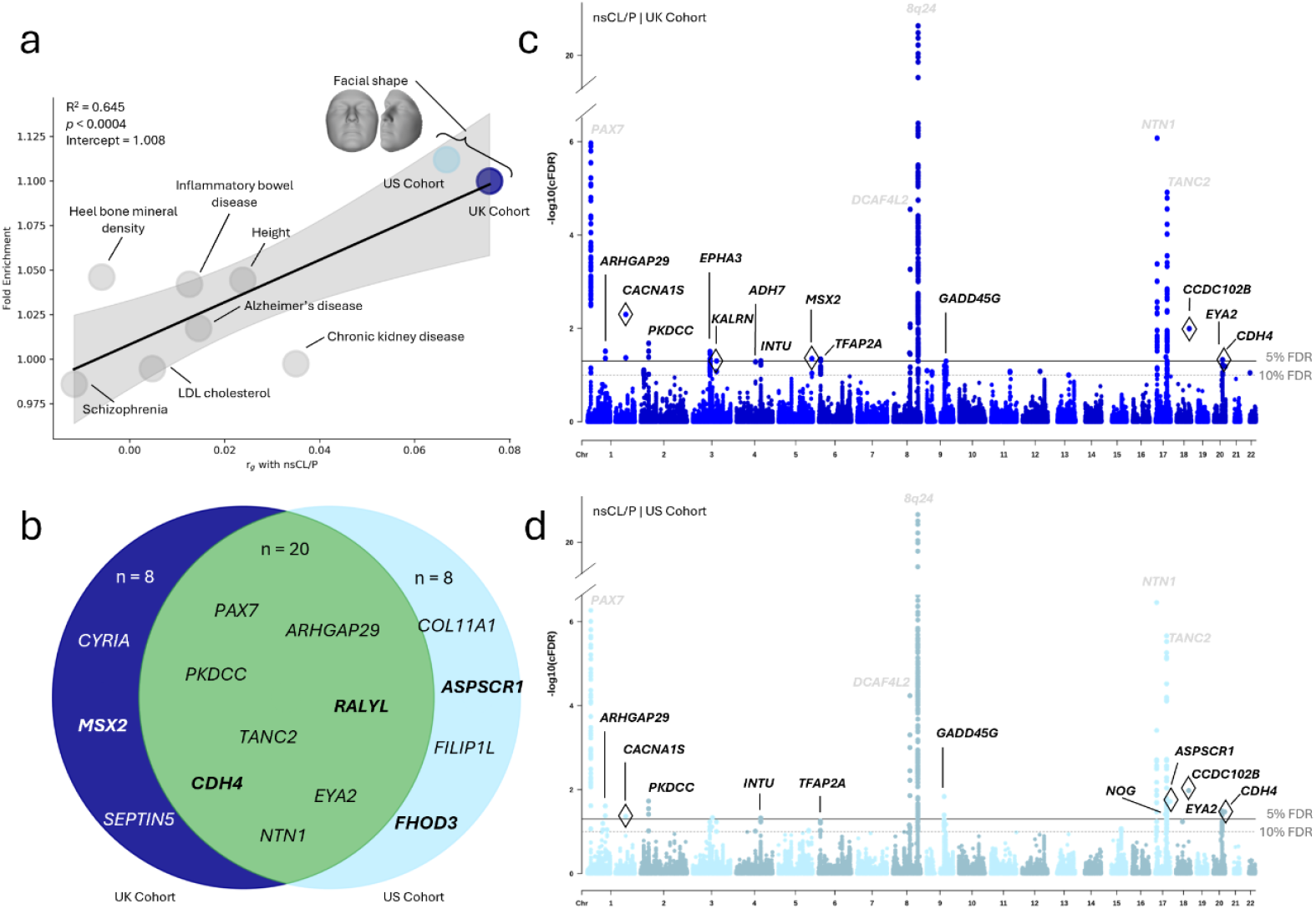
OFC loci enrichment and discovery results. **a)** The fold enrichment of nsCL/P SNPs (PnsCL/P< 0.05) in auxiliary traits’ SNPs (Pother< 0.05) compared to all SNPs with respect to each auxiliary traits’ genomic Spearman correlation with nsCL/P. GWAS sample sizes are as follows: nsCL/P (n= 3,969), schizophrenia (n= 36,989 cases and 113,075 controls), heel bone mineral density (n= 426,824), LDL cholesterol (n= 361,194), inflammatory bowel disease (n= 25,042 cases and 34,915 controls), Alzheimer’s disease (n= 71,880 cases and 383,378 controls), height (n= 2,200,007), chronic kidney disease (n= 64,164 cases and 561,055 controls), U.S. facial shape (light blue; n= 4,680), and U.K. facial shape (dark blue; n= 3,566). Iteratively reweighted least squares regression was used to estimate the trend line and 95% CI (P = 3.66e-4). **b)** A Venn-diagram highlighting selected known and new (bold) loci identified by the U.K. cohort (dark blue), U.S. cohort (light blue), and both (green). Lastly, there are two Manhattan plots illustrating the cFDR-GWAS results of nsCL/P | Face for the **c)** U.K. and **d)** U.S. cohorts. The solid horizontal line represents the 5% conditional FDR and the dashed line represents a suggestive threshold at 10% conditional false discovery rate. There is a y-axis break on both Manhattan plots to gain better resolution. Lead SNPs with diamond outlines represent new nsCL/P risk loci candidates.

### Pleiotropy-informed GWAS of nsCL/P enhances genetic discovery

We performed a pleiotropy-informed GWAS of nsCL/P using the conditional false discovery rate (cFDR) framework, referred to previously^35^ and hereafter as cFDR-GWAS. Specifically, we conditioned our large-scale GWAS meta-analysis of nsCL/P^12^ (Figure S1) on recent, large-scale GWASs of facial shape^36^ (denoted as nsCL/P | Face). Composite cFDR-GWAS results of nsCL/P | Face are shown in a Venn diagram (Figure 1b) with independent Manhattan plots for conditioning on two different facial shape GWAS datasets, one from the U.K. (Figure 1c) and one from the U.S. (Figure 1d). This approach of conditioning nsCL/P GWAS data on facial shape GWAS data boosted nsCL/P locus discovery. The cFDR approach identified 36 independent genomic loci associated with nsCL/P (Table S1), with 21 loci detected using a conditional FDR threshold of 5% and an additional 15 loci detected when the threshold was relaxed to 10%. To compare the number of detected loci with the original GWAS, we calculated FDR-adjusted P- values (defined by Benjamini and Hochberg)^37^ from the original nsCL/P GWAS (Figure S1). This revealed ten total signals at 5% and two additional signals at 10%, indicating that the pleiotropy-informed method retained 24 total signals that relied on leveraging the genetics of facial variation.

Effect sizes of the lead SNPs (absolute value of the logarithm of the odds ratios) are visualized for loci detected at 5% cFDR (Figure 2a) and 10% cFDR (Figure 2b) to demonstrate the comparable magnitude of effect at the two levels of identification. The majority of lead SNPs (55%) were detected in both the U.K. and U.S. cFDR-GWAS facial trait cohorts. However, there was a greater presence of association overlap between facial trait cohorts at the 5% cFDR threshold (86%) compared to 10% cFDR (13%). An approximately 60:40 ratio of known to new loci (39%) provided a unique opportunity to compare the effect sizes of new hits to known ones, within the context of a single study. All lead SNP effect sizes are greater than 0.2, with average magnitude of effect sizes of 0.31 and 0.35 for known and new loci, respectively (Figure 2c).

**Figure 2.**
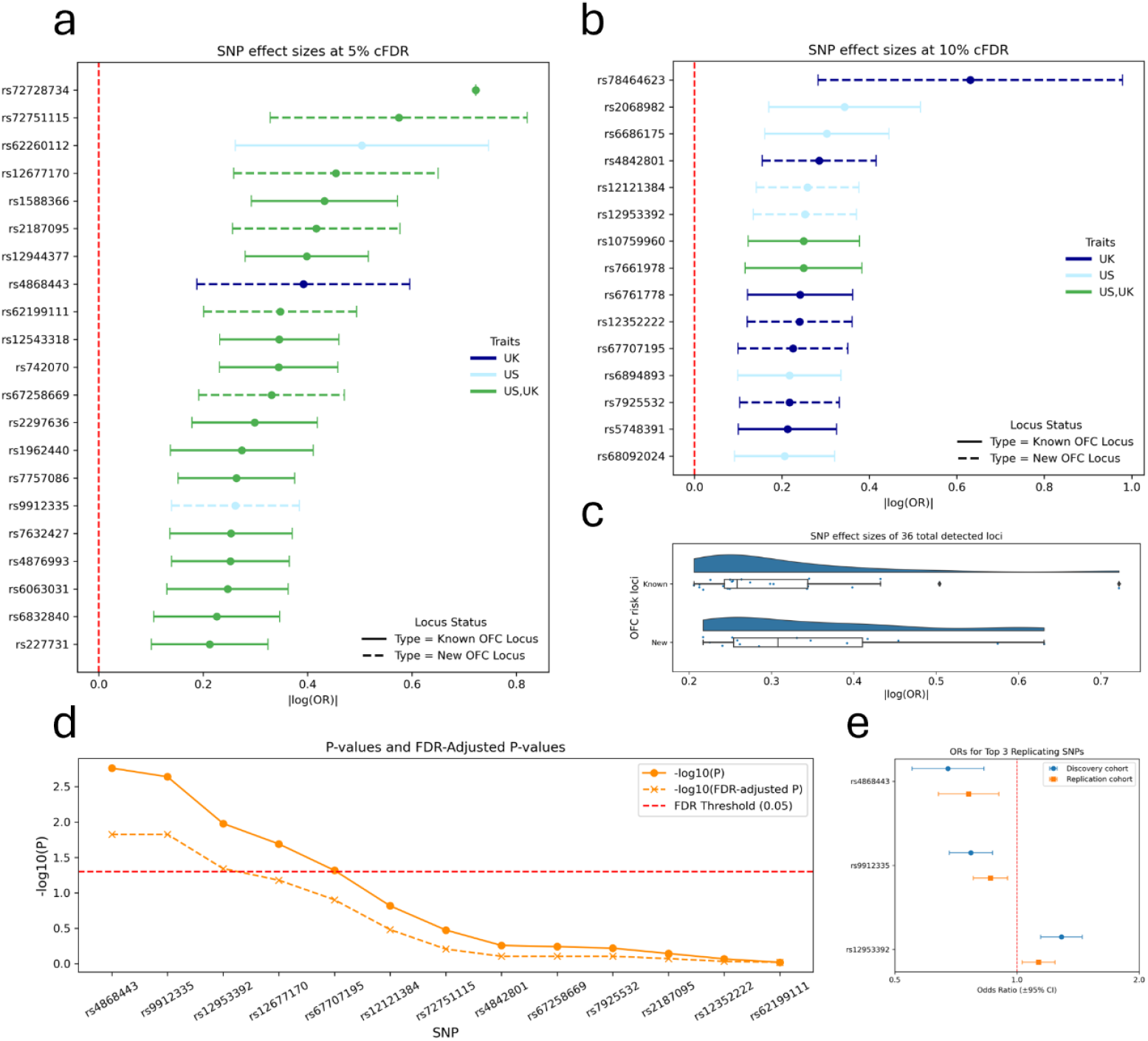
Validation of the 36 independent genomic loci detected. **a)** Forest plot of original nsCL/P GWAS effect sizes for the 21 OFC risk loci that were detected in the cFDR-GWAS approach at a cFDR threshold of 5% and **b)** the 15 OFC risk loci detected at 10% cFDR. Point and error bar colors represent which cFDR-GWAS the locus was detected (UK-dark blue; US-light blue; Both-green). The solid line style represents OFC risk loci that have already been identified and discussed in literature while the dashed line style represents the new loci that were detected in this current study. **c)** Comparison of original nsCL/P GWAS effect sizes between known and new OFC risk loci detected in the cFDR-GWAS approach. **d)** The new OFC risk loci identified in the cFDR-GWAS were subject to a replication lookup in an independent nsCL/P case-control cohort (orange) and their associations are represented by P-values (filled circle) and FDR-adjusted P-values (x). **e)** Forest plot of original odds ratios from the discovery nsCL/P GWAS (blue) and the replication nsCL/P GWAS (orange) for the top three new OFC risk loci that were replicated at an FDR threshold of 5%.

### Replication in an independent nsCL/P cohort

To validate the 14 new genomic loci, we tested each lead SNP for association in an independent nsCL/P case-control cohort^17^ of recent European ancestry (1,084 cases and 7,913 controls) from The Cleft Collective^38^. Of the 13 lead SNPs that were available for testing, we replicated five at the nominal significance level of p<0.05 (Figure 2d; Table S2). Although not every new locus replicated, we still observed more statistically significant signals than expected by chance alone (Binomial test p=2.9E-4). We also generated FDR values for the replication results which highlight lead SNPs at three loci that surpassed the 5% FDR threshold. These loci contained the candidate genes *MSX2, ASPSCR1*, and *FHOD3*. Our increased confidence in these top three externally validated signals motivated additional downstream analyses. Odds ratios for these top three replicating lead SNPs were obtained from the nsCL/P discovery and replication cohort GWASs (Figure 2e). All three lead SNPs showed consistency in risk versus protective alleles across the discovery and replication cohorts. Of note, these three signals would not have been detected in the original nsCL/P GWAS at a generalized FDR threshold of 5% (*Gene*:FDR adjusted P, *MSX2*: 0.50, *ASPSCR1*: 0.24, *FHOD3*: 0.24), reiterating the importance of the cFDR-GWAS framework and leveraging facial shape. Regional association plots were used to illustrate genetic associations of these three loci for the cFDR-GWAS, original nsCL/P GWAS, and original facial shape GWAS (Figure 3).

**Figure 3.**
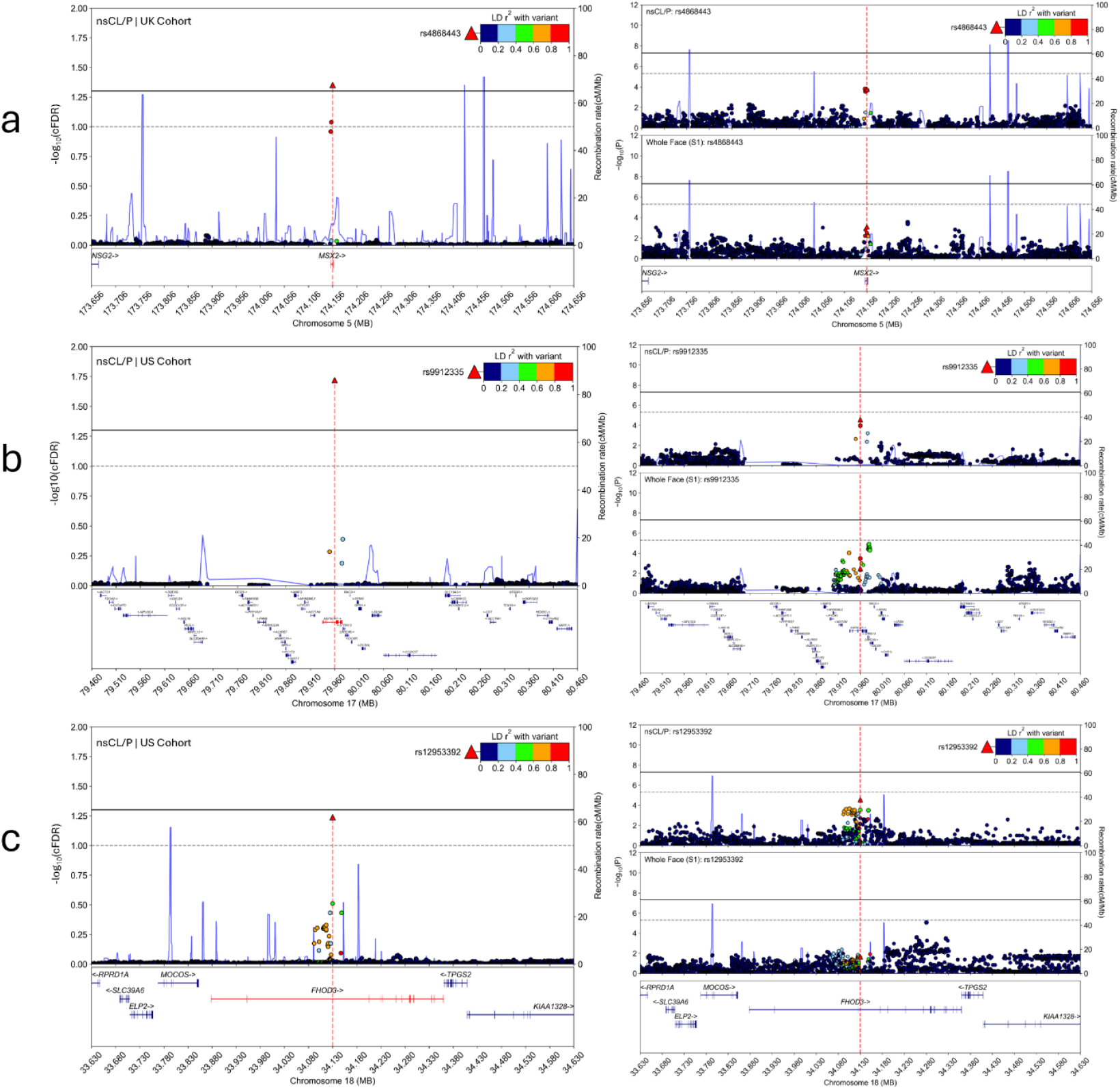
Regional association plots for the top three replicated loci near genes *MSX2, ASPSCR1*, and *FHOD3*, where the lead SNP is denoted as a red triangle for **a)** rs4868443, **b)** rs9912335, and **c)** rs12953392. Markers in linkage disequilibrium (LD) follow the color pattern in the top right legend of each plot. Each panel consists of three regional association plots that represent the cFDR-GWAS (left), original nsCL/P GWAS meta-analysis (top right), and original facial trait GWAS (bottom right). The black, solid line represents the significance threshold, either cFDR < 5% or P < 5e-8 (original GWASs). The gray, dashed line represents the suggestive threshold, either cFDR < 10% or P < 5e-6 (original GWASs).

### Cell-type specific gene expression and spatial transcriptomics

We turned to single-nucleus RNA sequencing (snRNA-seq) data to support the biological characterization of candidate genes at our three replicated OFC risk loci using a gene expression atlas of human craniofacial tissues^39,40^ during key time points of embryonic facial development. To understand the gene expression patterns of our top three replicating loci in CS13 human embryonic craniofacial tissue, we referenced the spatial transcriptomics data in the comprehensive atlas of craniofacial development^41^ (Figure 4) for genes at two known loci from the original nsCL/P GWAS (*PAX7* and *TANC2*) and the top three replicating loci identified here. Spatial transcriptomics and UMAP feature plots of main human cell types for the top three replicating loci are provided in Figure S2. Each of our top genes presents distinct patterns of gene expression. For example, *MSX2* exhibits localized, high gene expression in multiple regions including the pharyngeal arches, heart, somatic lateral plate mesoderm, and similar to *PAX7*, expression in the trunk neural crest. The spatial plot of *ASPSCR1* highlights expression in every cell cluster which is reflective yet more ubiquitous than the expression pattern observed in another known locus, *TANC2*. Our third replicating locus, *FHOD3*, presents with expression arranged primarily in the heart cell cluster, with traces of expression in the forebrain, craniofacial ectoderm, and trunk neural crest (Figure S2d).

**Figure 4.**
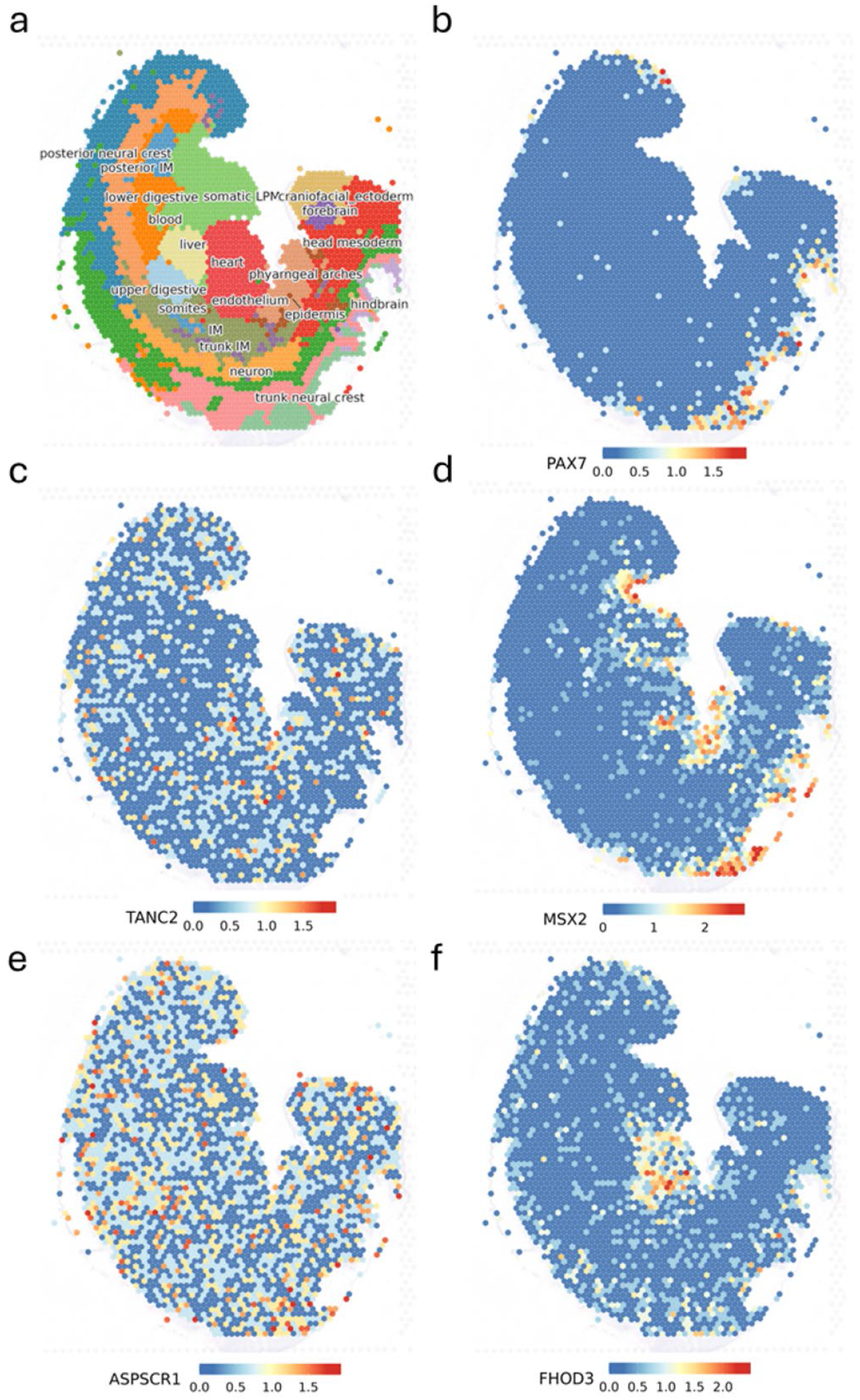
Gene expression at the cell-type level at Carnegie stage 13 of human development where panel **(a)** represents the spatial locations of each cell-type, panels for **b)** *PAX7* and **c)** *TANC2* represent gene expression coordinates of candidate genes at known loci discovered in the original nsCL/P GWAS, and the following panels **(d-f)** are specific gene expression patterns for nominated candidate genes (*MSX2, ASPSCR1*, and *FHOD3*) at the top three replicating loci in our study.

## DISCUSSION

In traditional GWAS, it has been shown that signals below the strict genome-wide significance threshold we impose by convention are enriched for true positives^42^. Analytical strategies that can disentangle these true positive signals from the complex landscape of sub-threshold associations are valuable, particularly when dealing with less common traits. In this study, we used the shared genetic architecture of OFCs and normal facial variation to detect several new OFC candidate risk loci. Like other congenital malformations characterized by complex genetics, a limitation of gene mapping studies on OFCs is their small sample size and the attendant low power to detect variants with moderate effect. Applying the conditional FDR approach allowed us to leverage many pleiotropic signals in the craniofacial complex connecting typical-range morphology (facial shape) and dysmorphology (OFCs). This is largely due to the empirical Bayesian framework, which mitigates the rate of false discovery by conditioning genetic associations with our trait of interest, nsCL/P, using prior evidence of SNP associations with facial traits. We addressed potential concerns that conditioning on any complex trait could improve detection of signals by illustrating significantly higher enrichments of SNPs in facial traits over several other auxiliary traits including height, low-density lipoprotein, and schizophrenia. Then, we performed conditional FDR analyses on nsCL/P given facial traits using summary-level P-value data from previous GWASs and identified 21 independent genomic loci at a 5% cFDR.

A major goal of studies on the genetic architecture of OFCs is to disentangle the complex and heterogeneous etiologies that give rise to all the OFC subtypes, and here we focused on nsCL/P. The main novelty is our application of a pleiotropy-informed GWAS method that leverages a genetically related trait to carefully control FDR and boost discovery. In our cFDR approach at significant and suggestive (0.05 and 0.10) thresholds, using SNP associations from facial shape as cFDR prior evidence, we detected 36 independent associated loci in total and should expect 10% (about 4 loci) of those results to be false positives. While 14 of our discovered signals showed no prior association with clefting, the inclusion of 17 loci previously implicated in OFC genetic studies, lends credence to the validity of the approach. Five of those new signals showed nominal evidence of association in a small replication cohort. Moreover, when we looked at odds ratios of known and new associations, we found similar magnitudes of effect sizes between these two classes of loci. This suggests that many of the newly discovered loci are not weaker signals uncovered by merely relaxing statistical thresholds. Rather, these reflect true associations that were previously undetected due to limited power but now visible by leveraging a closely related trait.

These findings contribute further evidence to a polygenic component of OFC risk, specifically nsCL/P, by revealing several associations that would have otherwise gone undetected. For example, the original nsCL/P GWAS used in this study, at a comparable suggestive threshold of detection (P < 5e-6), identified a fraction of the loci (11 out of 36) that were uncovered with the pleiotropy-informed GWAS approach, using the same nsCL/P GWAS summary statistics but leveraging the genetic relatedness of facial shape. The new candidate loci nominated here add to the growing network of previously reported loci associated with OFCs, offering new insights into other sets of genes that contribute to OFC risk while providing evidence of pleiotropic loci given their enrichment in other biological processes and systems. More generally, this approach offers a scalable way to interrogate OFC genetics across diverse study designs and cohorts while potentially providing a more nuanced view of the underlying genetic architecture.

As previously mentioned, 5 of 13 (38%) new OFC risk loci candidates replicated in an independent European cohort of 1,084 nsCL/P cases and 7,913 controls at a nominal significance threshold with P-values less than 0.05. Not every new signal replicated, but a replication rate of 38% suggests more frequent associations than chance alone. When we applied a more stringent significance threshold using 5% FDR in the SNP candidate test, we detected three loci implicating the genes *MSX2, ASPSCR1*, and *FHOD3*, making these three genes among the most robust candidates from the new loci identified in our genome-wide study.

*MSX2, ASPSCR1*, and *FHOD3* represent biologically distinct OFC risk candidates emerging from the polygenic background likely responsible for moderate involvement in OFCs. Indeed, the *MSX2* gene is already a putative candidate for involvement in OFC, however, it has eluded associations at the level of genome-wide significance, appearing in a gene-candidate test^43^ or being in proximity of a suggestive hit^44^. Of note, the suggestive hit was described as having some evidence for association with cleft lip and palate (CLP), specifically, and identified through genome modifier tests^44^. However, all eight SNPs reported are located in a long intergenic non-coding RNA (*LINC01411*), are not in linkage disequilibrium with our lead SNP (r^2^ = 0), and are located 250 kb or more away (closest SNP is rs72820767, which is 248 kb upstream). Importantly, *MSX2* is well known for its link with Boston type craniosynostosis^45-47^ and has established molecular mechanisms^48,49.^ The co-implication of common variants in craniosynostosis and OFC has been reported previously^50^, and the present finding aligns with this surfacing pattern of shared craniofacial genetic risk.

*ASPSCR1* resides on chromosome 17 (17q25.3) and encodes a protein that interacts with glucose transporter type 4 (GLUT4)^51,52.^ There is some evidence this pathway may be relevant to craniofacial development as *TRARG1*, a trafficking regulator of GLUT4, is associated with the nsCL/P endophenotypic (subclinical) facial morphology of the philtrum^26^. Although direct mechanistic links remain unclear for our candidate gene, *ASPSCR1*, recent developmental evidence has shown that proper glucose uptake in cranial neural crest cells (CNCCs) is required for midfacial morphogenesis, with disruption of this metabolic–epigenetic pathway leading to midline facial clefts^53^.

Finally, our third replicating signal in *FHOD3* indicates another diverse yet plausible shared pathway between the face and the heart. The spatial transcriptomics (Figure 4f; Figure S2d) of this gene highlight preferential expression in heart cell types at CS13 with small to moderate expression in the forebrain, craniofacial ectoderm, and trunk neural crest. It is a known disease-causing gene in hypertrophic cardiomyopathy^54^ (HCM) which is characterized by asymmetric septal hypertrophy, an unevenly thickened septum. This is potentially relevant because congenital heart defects are more common in individuals with OFC than the general population with an incidence rate at approximately 15%^55,56^.

Orofacial clefting is considered etiologically heterogeneous and may involve diverse genetic pathways and/or biological processes. A long standing hypothesis holds that the shape of the embryonic face may itself be a risk factor for OFCs^27^. The basic premise is that any factor that alters the tightly coordinated arrangement among the rapidly developing adjacent tissue masses that form the face can impact their ability to meet and fuse, thereby increasing the chances of cleft. Changes in the relative spatial arrangement of those tissue masses (i.e., embryonic facial shape) is hypothesized to be one of those predisposing factors. The relationship between OFC and facial shape is supported by several converging lines of evidence: strains of mice at high risk for clefting show morphological differences in their faces as embryos and adults compared to low risk strains^57,58^; the parents and siblings of individuals affected with clefts show facial shape differences compared to the general population^26^; GWAS of typical-range facial shape show a statistical overrepresentation of genes associated with OFC^24^; and previously identified cleft risk variants have been shown to impact human facial shape in predictable ways^59,60.^ Our results provide additional strong support that the relationship between human facial shape and OFC is driven by a partly shared genetic architecture.

The genetics of OFCs have been investigated for decades with myriad studies exploring many different independent datasets, each contributing information into the complex genetic heterogeneity and etiology of the birth defect. An open question is whether the collection of genes identified here (or a subset of them) contribute to a distinct shape-driven etiological pathway capable of influencing OFC liability. This could be conceptualized as a polygenic background effect, pushing individuals toward or away from the liability threshold. Such a background effect could operate epistatically by modifying risk or phenotypic severity in the presence of one or a few genes with more pronounced effects. Yu et al. (2022) for example showed that polygenic background effects (modeled as a risk score comprising common variants) can modify the phenotypic penetrance of rare damaging mutations in nsCL/P^61^. Thus, the work here may provide a foundation for modeling penetrance and predicting phenotypic severity. Furthermore, this approach may hold similar promise for other complex traits and structural birth defects.

## METHODS

### GWAS summary-level source data

Genetic association results were recalculated with one degree of freedom from our large 2017 genome-wide meta-analysis of nsCL/P run in a subset of 3,969 individuals of recent European genetic ancestry^12^. Using one degree of freedom was necessary to avoid the deflation observed in the original analysis (Figure S1). For facial shape (the main conditioning trait), we used summary statistics from our 2021 GWAS involving 4,680 United States (U.S.) individuals and 3,566 United Kingdom (U.K.) individuals, each of recent genetic European ancestry^24^. These facial shape analyses were based on a 3D facial surface divided into 63 segments using a data-driven approach^23^ and multivariate GWAS was then run on measures of shape for each segment separately. Using that approach we identified 203 distinct genome-wide significant signals at 138 loci associated with facial variation. For the present study, we conditioned on the primary, whole-face GWAS (facial segment 1) and we did this for the U.S. and U.K. cohorts separately to compare the outcomes.

In addition, we conditioned using the summary statistics for other recent large-scale GWAS studies of complex traits with no known link to OFCs to determine how these would perform compared to facial shape. These traits included schizophrenia^28^, heel bone mineral density, low-density lipoprotein cholesterol^30^, inflammatory bowel disease, Alzheimer’s disease, height^33^, and chronic kidney disease and the GWAS summary statistics we used were from analyses of cohorts of recent genetic European ancestry.

### Genomic Spearman correlations

To determine if nsCL/P is more genetically correlated with facial traits than several other complex traits mentioned above, we performed genomic spearman correlation analyses. The summary statistics of our primary trait of interest, nsCL/P, were derived from modelling a binary trait in a univariate GWAS and support traditional genomic correlation calculation via linkage disequilibrium (LD) score regression^62^. However, our closely related secondary trait of interest, facial shape, provided summary statistics from a continuous trait modelled in a multivariate GWAS, which does not permit the traditional genomic correlation approach.

Therefore, following an approach with a similar challenge^63^, we calculated genomic Spearman correlations across independent LD blocks^64^ (excluding the major histocompatibility region: GRCh37 positions of chr6:25,119,106–33,854,733) estimated in European populations^65^ since our GWAS summary data were all derived from analyses of individuals with predominantly genetic European ancestry. The calculation for the genomic Spearman correlation (python 3.11, spearmanr from scipy.stats) between traits was based on previous work^35^.

### Cross-trait enrichment of statistical association

Next, we attempted to determine the degree to which nsCL/P was genetically enriched in facial traits compared to the auxiliary complex traits mentioned above. Enrichment was evaluated by comparing SNPs with statistical associations to nsCL/P to SNPs associated with another trait. Enrichment was indicated by a fold increase in the fraction of nsCL/P associated SNPs (P_nsCL/P_ < 0.05) among secondary trait associated SNPs (P_Other_ < 0.05) relative to the set of all SNPs^35^. The increase in tail-probabilities in the P-value distributions of the fraction of SNPs relative to the full set provides a measure of the fold-enrichment.

### Conditional FDR analysis

The conditional FDR approach was first introduced by Andreassen^19^ to study psychiatric traits and was recently applied by our team to investigate craniofacial phenotypes^35^. Briefly, we define this as the probability that a SNP has no true association with the target trait (nsCL/P), given that the observed P-values for both the target and a related trait (facial shape) are as small or smaller than expected. In other words, this work is based on a generalized empirical Bayesian view of FDR originally proposed by Efron^21^.

Conditional FDR analysis was performed using the pleioFDR software^19^ (https://github.com/precimed/pleiofdr). To minimize bias from regions of complex LD, we excluded SNPs located within the major histocompatibility complex on chromosome 6 and the 8p23.1 region (GRCh37: chr6:25,119,106–33,854,733 and chr8:7,200,000–12,500,000). Final models were built using 500 iterations of LD-pruned SNP sets, where pruning was performed on SNPs at random and at a threshold of r^2^ < 0.2.

### Identification of independent genomic loci

The independent loci from the nsCL/P cFDR-GWAS framework were identified using default settings for FUMA^66^ v1.5.2. In total, there was one cFDR-GWAS per facial shape cohort. Independent lead SNPs were first identified using LD estimates from European samples in the 1000 Genomes Project Phase 3 dataset^65^, applying an r^2^ threshold of 0.1. A final set of independent loci across both cFDR-GWAS analyses (U.S. and U.K. cohorts) was generated through a merging process based on a ±250 kb distance criterion. Lead SNPs located within 250 kb of each other were grouped into a single locus, with the most significant SNP representing the locus. Throughout the manuscript, the term “lead SNP” refers specifically to this representative SNP at each independent locus.

### Replication in an independent nsCL/P cohort

Lead SNPs that have not previously been associated with OFCs were tested for association in an independent nsCL/P cohort of recent European ancestry. The summary statistics that were used for the replication analysis represent a dataset from The Cleft Collective^38^, a longitudinal cohort study of children born with CL/P and their families (https://www.bristol.ac.uk/cleft-collective/). It includes 1,084 nsCL/P cases and 7,913 controls recently analyzed by Dack et al.^17^ (2025). Access to this data is also described in the data availability section.

### Gene annotation

For each identified locus, no more than two candidate genes were assigned using GREAT v4.0.4 (default settings, binomial test) by selecting those with the nearest transcription start sites (TSS) on either side of the lead SNP, as long as their TSSs were situated within 1 Mb or overlapped curated regulatory elements. If there were no genes that could be clearly implicated, we manually selected genes that were within 1 Mb of the lead SNP, strongly supported by existing, relevant research. Additionally, these manually annotated genes were required to be in the same topologically associating domain (TAD) based on craniofacial combined HiC maps found on the UCSC Genome browser.

### Gene expression patterns by cell type in craniofacial development

To illustrate the biological relevance of our findings, the genes assigned to replicated lead SNPs were subject to gene expression visualization tools using the interactive web application provided by the comprehensive atlas of human craniofacial development^41^. Using these tools, we generated gene expression plots for major cell types and subtypes of embryos at timepoints relevant to facial development, including spatially located cell type-specific coordinates on human (CS13) spatial transcriptomic data^39,40.^ Additional information about the human craniofacial atlas and web application is found in Khouri-Farah et al. (2025).

## DECLARATIONS

### Data Availability Statement

The facial shape GWAS summary statistics from White et al. (2021; GCST90007181; GCST90007244) and European nsCL/P summary statistics from Leslie et al. (2017; GCST90652505) are available through GWAS Catalog. The case-control nsCL/P cohort used for replication can be obtained from The Cleft Collective by request or summary statistics can be found on GWAS Catalog (GCST90709866). For snRNA-seq gene expression data, code for analysis and generating figures is found on github (https://github.com/cotneylab/craniofacial_snrna). An interactive website for exploring processed data is found here: https://cotneyshiny.research.chop.edu/shiny-apps/craniofacial_all_snRNA/.

### Ethical Oversight

The main cFDR analysis uses publicly available summary statistics from previously published work. All participants provided their written informed consent or assent to prior to participation in any research activities. The use of human embryonic tissue was reviewed and approved by the Human Subjects Protection Program at UConn Health (UCHC 710-2-13-14-03) and Children’s Hospital of Philadelphia (IRB 24–022258). Human embryonic craniofacial tissues were collected via the Joint MRC/Wellcome Trust Human Developmental Biology Resource (HDBR) under-informed ethical consent with Research Tissue Bank ethical approval (18/LO/0822 and 18/NE/0290, project 200225). Donations of tissue to HDBR are made entirely voluntarily by women undergoing termination of pregnancy. Donors are asked to give explicit written consent for the fetal material to be collected, and only after they have been counseled about the termination of their pregnancy. Further documentation of all policies and ethical approvals for HDBR sample collection can be found at https://www.hdbr.org/ethical-approvals.

### CREDiT

Conceptualization: SMW, JRS, PC, NH, SG; Data curation: NH, AM, MKL, XZ, JCC, AD; Formal analysis: NH; Funding acquisition: SMW, JRS, MLM, JC; Investigation: NH, SMW, JRS; Methodology: NH, SMW, JRS, SG, PC; Project administration:; Resources: SMW, JRS, MLM, PC, JC, EJL, JCC, SJL; Software:; Supervision: SMW, JRS; Validation: NH; Visualization: NH, SG, AM, JC; Writing – original draft: NH, SMW, JRS; Writing – review & editing: NH, SMW, JRS, MLM, SG, PC, AM, JC, EJL, JCC

