## Supplementary Figures for "Leveraging the genetics of human face shape boosts the discovery of orofacial cleft risk loci"

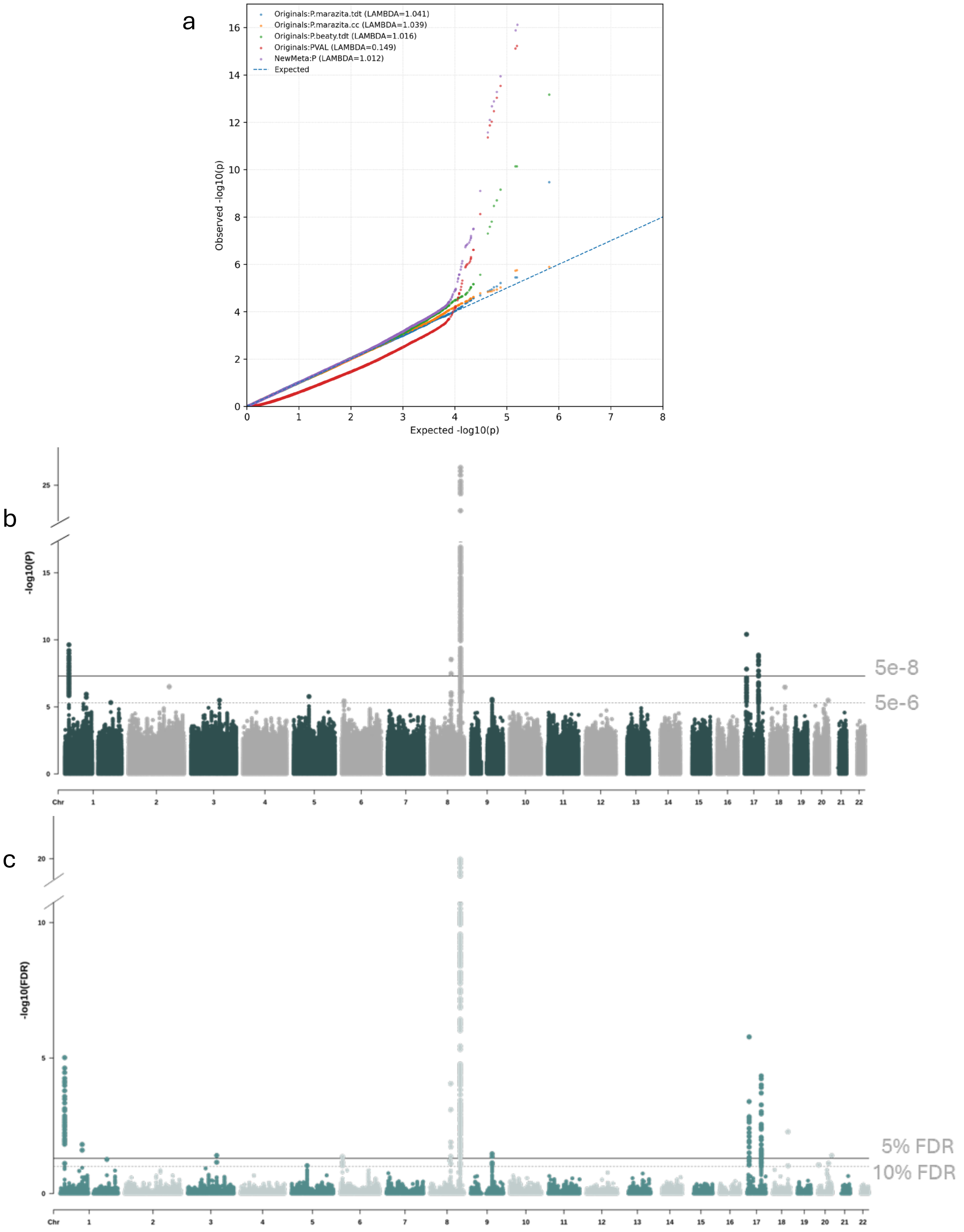


**Supplementary Figure 1** Genome-wide association results for the original nsCL/P discovery cohort. **a)** QQ-plot of the three GWAS studies (blue, orange, and green) used for the meta-analysis reported in 2017 (red). The same meta-analysis was performed with one degree of freedom to avoid unnecessary deflation (purple). **b)** Manhattan plot of the recalculated, original genome-wide meta-analysis of nsCL/P run in a subset of 3,969 individuals of recent European genetic ancestry with genome-wide significance threshold at 5e-8 and suggestive threshold at 5e-6 and **b)** the FDR values of the same associations with thresholds at 5% and 10% FDR.

**Supplementary Figure 2** Gene expression feature plots per gene for the top three replicating loci demonstrating normalized levels of expression in UMAP dimension reduction plots, CS13 human embryo spatial plot section one, and CS13 human embryo spatial plot section two. Each figure has the map of cell-type (UMAP) and subtype (spatial plots) clusters displayed in the top row of the figure for reference. The following figures and panels are: **a)** reference map, **b)** *MSX2,* **c)** *ASPSCR1,* and **d)** *FHOD3.*

Fig. S2

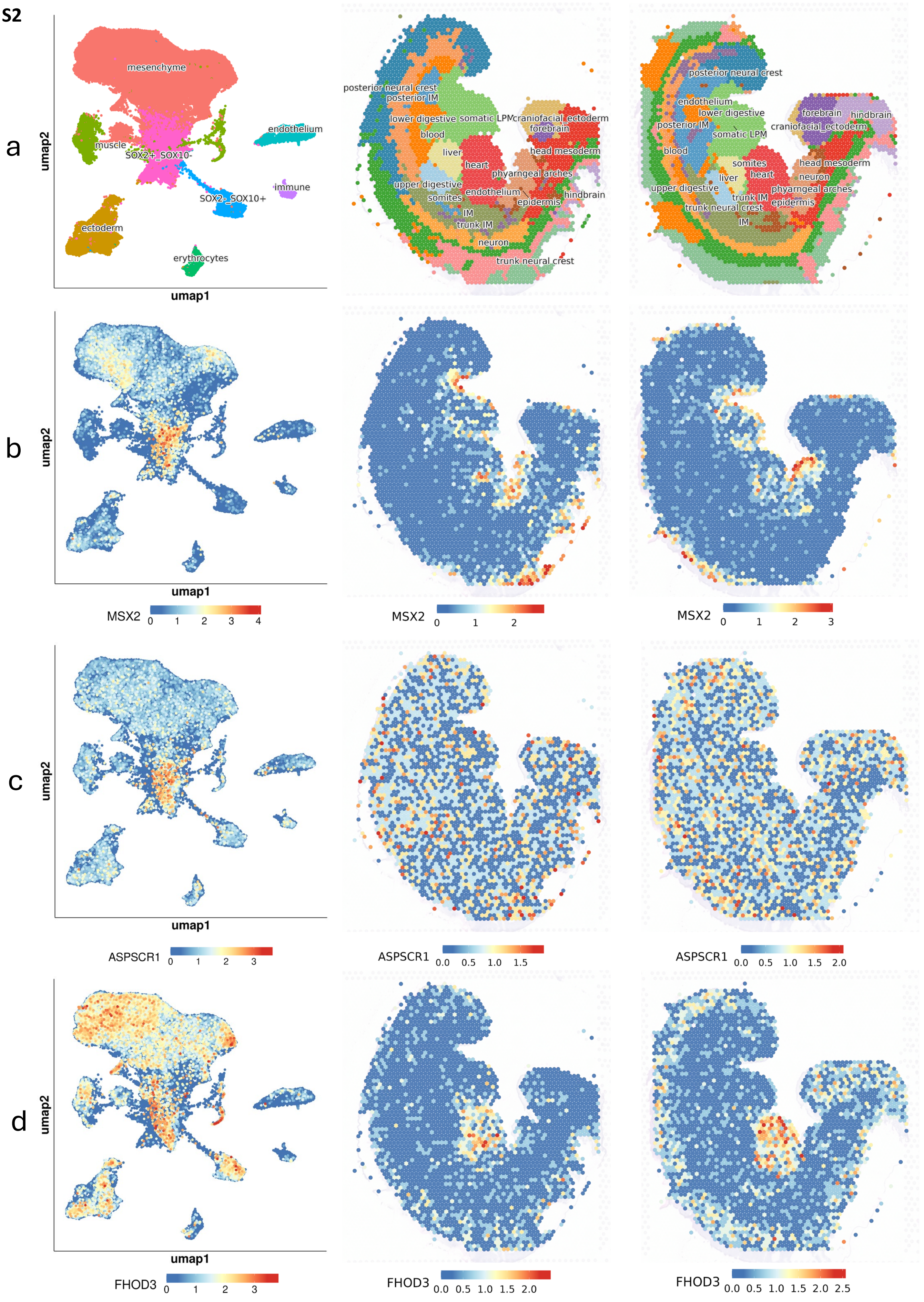
a
b
c
d
umap1
umap2
mesenchyme
muscle
SOX2+_SOX10-
endothelium
immune
SOX2-_SOX10+
ectoderm
erythrocytes
posterior neural crest
posterior IM
lower digestive
somatic LPM
craniofacial_ectoderm
forebrain
blood
liver
head mesoderm
heart
phyarngeal arches
upper digestive
endothelium
somites
IM
epidermis
hindbrain
trunk IM
neuron
trunk neural crest
MSX2
ASPSCR1
FHOD3
